# Development and validation of a real-time PCR assay for the diagnosis of rabies virus Philippine strain in non-brain samples

**DOI:** 10.1101/2024.12.04.24318476

**Authors:** Daria L. Manalo, Jude Karlo G. Bolivar, Jeromir G. Bondoc, Blanca J. Nagataki, Leilanie B. Nacion, Mark Joseph M. Espino, Chun-Ho Park, Satoshi Inoue

## Abstract

Rabies is a fatal neurotropic and zoonotic disease responsible for thousands of deaths yearly. Direct fluorescent antibody test (dFAT), the gold standard in routine rabies diagnosis, requires dog brain samples, and takes 5-7 hours to obtain results. Brain specimen degradation due to inappropriate transport and storage conditions most of the time leads to false negative results, hence the need for an alternative diagnostic method that can also utilize non-brain specimens. In this study, an RT-qPCR assay was developed to specifically target the N gene of the Philippine rabies isolate. The assay was optimized using RNA from dFAT-positive dog brain tissues as templates. In-silico and in vitro evaluations both showed 100% specificity to rabies RNA, with a detection limit of 1 copy per microliter. Validation of the assay was done using dFAT-tested brain samples and potential brain specimen-alternates, specifically the dog nasal planum (NP) and follicle sinus complex (FSC). One hundred percent of the NP and FSC samples showed concordance with the respective dFAT-positive brain samples. Only 97% concordance was observed with the dFAT-negative brain samples. These results collectively validate the efficiency, sensitivity and specificity of the assay developed, indicating its potential utilization for in rabies diagnosis using clinical samples besides the brain tissues.

## INTRODUCTION

Rabies is a zoonotic disease that affects the central nervous system (CNS) ultimately causing death if immediate post exposure prophylaxis (PEP) is not given. Infected dogs account for up to 99% of human rabies cases. The virus is primarily transmitted to humans and animals through bites and non-bite exposures, such as scratches, abrasions, or open wounds exposed to the saliva or other potentially infectious material from an infected animal (Rupprecht, 1996; CDC, 2019; CDC, 2021; WHO, 2021). Although a vaccine preventable disease, rabies remains a neglected and remarkably underreported disease posing a serious public health and economic burden in developing countries. The disease kills over 59,000 people around the world annually, resulting in an estimated global economic burden of USD 8.6 billion per year (Hampson *et al*., 2015).

The Philippines, being one of the major endemic countries in Southeast Asia, records nearly 200–300 human rabies cases annually (DOH, 2020). In 2007, the government-initiated Republic Act No. 9482, also known as the Anti-Rabies Act of 2007, to impede and curb human rabies in the country. However, between 2006 and 2015, San Lazaro Hospital, a sub-national laboratory in the Philippines, consistently admitted human rabies patients with no discernible decrease in the number of cases during that period. Most of the clinical features were shared by patients who were admitted between 1987 and 2006 and regions with the highest frequency of human rabies cases remained unchanged as well (Guzman et al., 2022).

Early detection and treatment of rabies infection lowers the mortality rate of both human and animal rabies victims. At present, the gold standard and most prevalent method used for the detection of rabies in animals is the direct fluorescent antibody test (dFAT) which requires brain tissues to detect rabies antigen.

Although this technique provides fast and accurate results, its sensitivity decreases and becomes unacceptable with the decomposition of the brain (WHO, 2018). This is particularly a concern in areas with elevated temperatures which can affect the transport of brain samples, leaving them unfit for laboratory examination. In tropical countries like the Philippines, with temperature ranging from 29 to 35 °C during the summer, decomposition of the brain is relatively faster. In a mouse experiment, the time-point limit of detection of antigen in the brain using dFAT was only 3 days when carcass was stored at 35°C, and beyond this, rabies antigen was undetectable (McElhinney *et al*., 2014). In contrast, viable RNA from naturally positive brain samples stored at 40°C was still detectable until 36 days (David *et al*., 2002).

Molecular approaches have recently been explored in diagnostics as they offer numerous benefits over conventional approaches, including improved sensitivity, higher throughput, and the potential for variant typing. Reverse transcription PCT (RT-PCR) tests, one of these emerging diagnostic technologies, enable the detection of rabies virus in tissues with low viral loads, such as saliva, nuchal skin biopsies, and corneal swab (Maier *et al*., 2010). According to Duong *et al*. (2016), PCR-based assays may be used as a confirmatory assay in parallel to other rabies diagnostic tests such as dFAT and ELISA, both in antemortem and postmortem diagnosis. These tests are highly sensitive but conventional gel-based RT-PCR assays are susceptible to cross contamination and non-specific amplification, which can lead to false-positive results. Specificity of the test can be improved by sequencing the PCR products. In comparison, reverse transcription real-time PCR (RT-qPCR) tests have increased sensitivity and specificity, demonstrating its potential to take the lead in the field of rabies laboratory diagnosis (Duong et al., 2016). The most common and widely used RT-PCR assay for rabies diagnosis is LN34 Pan-Lyssavirus assay which targets a highly conserved region of the nucleoprotein (N) gene and can detect a wide range of lyssaviruses across different species. Even in deteriorated and formalin fixed samples LN34 primers can be used in either conventional or qPCR assays with high diagnostic sensitivity and specificity for rabies virus detection (Gigante *et al*., 2018).

In 2006, two rabies cases were reported in Japan after developing infection 2-3 months after dog-bite exposure in the Philippines. A cocktail of primers was utilized to amplify the nucleoprotein-coding region of the rabies virus, and the amplicons were then subjected to sequencing. The acquired sequence was thereafter used as template to generate primers and probes specific for the virus strain isolated from the Philippines (Tobiume et al., 2009). In this study, these primers and probes were further utilized to develop an RT-qPCR assay to detect the presence of rabies RNA in dFAT-tested brain samples. The assay was thereafter validated using non-brain samples such as follicle sinus complexes (FSC), and nasal planum (NP). The assay may be valuable in detecting rabies virus particularly the strains specific to the Philippines and subsequently be utilized to enhance rabies diagnosis and surveillance in the country.

## METHODS

### 1. Ethical and biosafety considerations

A biosafety clearance was obtained from the Biorisk Management Office of the Research Institute for Tropical Medicine (RITM) allowing the utilization of the study protocol. An Institutional Animal Care and Use Committee certification was deemed unnecessary because there were no live animals used in the duration of this study.

### 2. Sample collection

Brain, follicle sinus complexes and nasal planum samples submitted to RITM and those collected from Regional Animal Disease Diagnostics Laboratory (RADDL) were used in this study. These samples were stored in −80°C freezer until further processing.

#### 2.1. Brain collection

Dog heads were prosected along the skull, to reveal the brain. Skull cap was removed by skinning the top of the head from behind the poll to the level of the eye. A knife was then used to puncture and cut through the underside. Skull cap was removed with four cuts made with a hatchet and mallet that are slightly angled inward. The dura, which covers the brain and lies between the left and right hemispheres, was cut to separate the cerebrum from the cerebellum. Hippocampus, cerebellum, and brainstem were collected because these areas have a relatively higher localization of rabies virus.

#### 2.2. Follicle Sinus Complex (FSC) collection

An incision around the hair bulb of the sinus was made, revealing the follicle sinus. Once the follicle sinus is visible, a transverse section was cut at the level of the ring sinus of the FSCs.

#### 2.3. Nasal Planum (NP) collection

An incision along the midline of the front of the dog’s nose was made that run as far back as possible. The transversal aspect of the nose was cut at the level of the alar fold at about 5mm thick excess tissue was removed, and the nasal planum (NP) was collected.

### 3. RNA Extraction

Thirty milligrams of tissue samples were added to 500µL lysis buffer in a bead beating tube (BMS 2641-0B) containing a one-piece 5mm stainless steel bead (BMS SS50-0001). Using the Bertin Precellys Tissue Homogenizer (Bertin Technologies, France), the tubes were subjected to four cycles of homogenization at 8000 rpm for 30 seconds. Incubation in a cooling rack (0°C to −20°C) for 50 seconds immediately followed at the end of each beating cycle. RNA was then extracted using the RNeasy Mini kit (Qiagen, Germany) according to the manufacturer’s instructions. The extracted RNA was eluted in 50 µL RNase-free water and stored at −80 °C until use.

### 4. Controls

Rabies viral RNA were extracted from dFAT-positive brain tissue samples and amplified by qRT-PCR targeting only the rabies N-gene. The qRT-PCR products were then pooled, and subsequently purified using QIAquick PCR Purification Kit (Qiagen, Germany). The purified products were used as positive control. One thousand copies of purified Rabies N gene per microliter (10^4^) was used as the concentration of the positive control. Nuclease free water served as the template for no template controls.

### 5. Primers and Probe

The RT-qPCR assay for rabies used primers and probes specifically designed to target rabies N gene. They were generated based on the sequence of rabies virus in autopsy cases in Japan caused by a strain from the Philippines (Tobiume et al., 2009). The sequences of these primers and probe are indicated in Table 1.

**Table 1.**
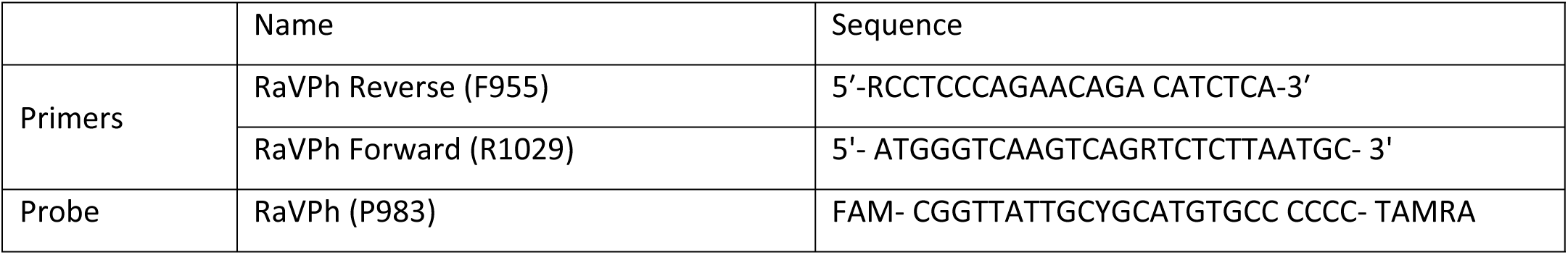
Primers and probe sequences.

### 6. Optimization of qPCR conditions

Multiple parameters were considered to obtain the optimal reaction conditions. These include primers and probe concentrations, primer annealing temperature, primer stability, assay specificity, assay limit of detection, and assay repeatability. One-step probe-based real-time PCR assay was initially employed following the recommended volumes (Table 2) and cycling conditions from the iTaq Universal Probes One-Step Kit (BioRad Laboratories, USA) with BioRad CFX96 Touch (BioRad Laboratories, USA) machine as platform. All parameters were tested using positive control and a no template control.

**Table 2.** RT-qPCR master mix preparation.

| PCR Component | Volume (1X) (in μL) |
| --- | --- |
| iTaq Universal Probes One-Step Kit | 10.0 |
| Forward primer (6.0μM) | 1.0 |
| Reverse primer (6.0μM) | 1.0 |
| Probe (3.0μM) | 0.50 |
| iScript™ reverse transcriptase | 0.50 |
| Nuclease free water | 4.0 |
| Nucleic Acid Template | 3.0 |
| TOTAL | 20 |

#### 6.1. Optimization of the annealing temperature

The optimal annealing temperature was determined using a temperature gradient ranging from 56°C to 65 °C. Specific temperatures within the gradient were pre-determined by the CFX PCR machine (56.8 °C, 57.3 °C, 58.4°C, 60.0 °C, 61.9 °C, 63.5 °C, 64.4 °C and 64.8 °C). The optimal temperature was determined as the one which exhibited the lowest Ct value without non-specific amplifications.

#### 6.2. Optimization of primers and probe concentrations

A primer concentration gradient (10.0µM, 8.0µM, 6.0µM, 4.0µM and 2.0µM) from varying dilutions of the stock primers (100µM) was done to determine the optimal primer concentration for the assay. The lowest working concentration from the gradient is assigned as the optimal primer concentration. The same method was employed to determine the optimal probe concentration (0.50µM, 1.0µM, 2.0µM and 3.0µM). After determining the optimal primer and probe concentrations, primer and probe stability was tested by running one positive control against 20 no template controls (NTC) using the optimized annealing temperature.

#### 6.3. Determination of assay efficiency, sensitivity, and specificity

The optimized thermocycling conditions in Table 3 were used in the subsequent laboratory performance validation of the assay.

**Table 3.**
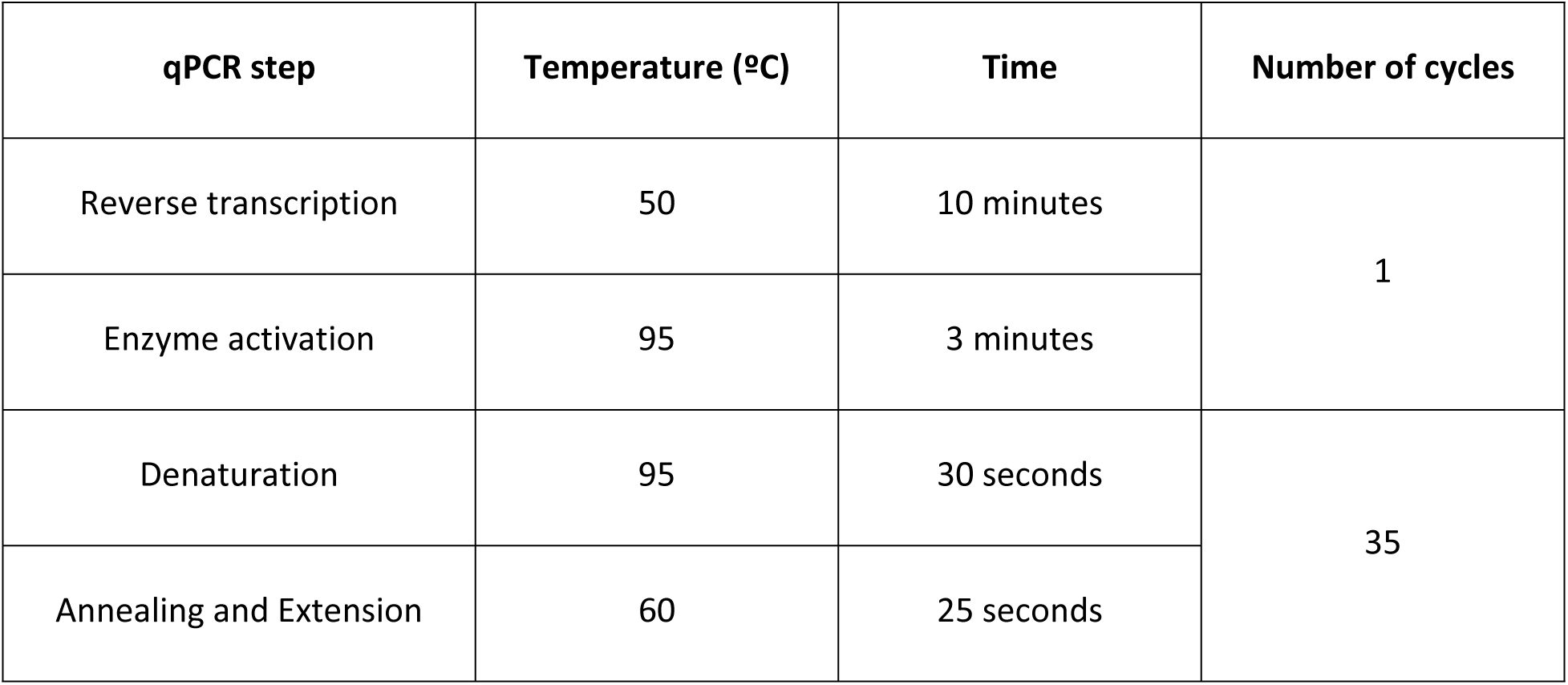
RT-qPCR profile for rabies N gene detection.

To determine the assay efficiency, a standard curve using dilutions of the positive control was established. The dilutions were as follows: 10⁶, 10^5^, 10^4^, 10^3^, 10^2^, 10^1^ and 10^0^. One PCR reaction mix of each template dilution was prepared and tested using the optimized thermocycling conditions.

To determine the assay’s limit of detection, a serial dilution (10⁶, 10⁴, 10^3^, 10^2^, 10^1^, 10^0^, 10^-1^, 10^-2^ and 10^-3)^ of the positive control was run with eight replicates each. The lowest dilution with a detection rate of 8/8 is determined as the limit of detection.

To evaluate the clinical specificity of the assay, positive control, canine parvovirus, and canine distemper virus were run using the optimized protocol. All samples were run in triplicates.

#### 6.4. Assay reproducibility

Assay reproducibility was tested by running the same protocol in different PCR machines and by different personnel within one week after the optimization. At least three different personnel performed the same assay in three different PCR machines.

#### 6.5. Assay validation and verification

The developed RT-qPCR assay was validated using dFAT-confirmed samples. Twenty dFAT-positive and thirty dFAT-negative samples were selected and extracted to serve as templates for the PCR run. A verification test to determine the Positive Percent Agreement (PPA) and Negative Percent Agreement of the assay to non-brain samples was done by using the FSC and NP corresponding to the dFAT-confirmed positive ang negative brain samples.

## Results

An RT-qPCR assay for the detection of rabies targeting the N gene was developed and optimal assay conditions were determined. The optimal annealing temperature is 60°C, showing the lowest Ct value without non-specific amplification (Table 4). Late, non-specific amplifications on non-rabies clinical samples were observed on annealing temperatures lower than 60°C. On the other hand, the optimal primers and probe final concentrations were 0.3µM and 0.075µM, respectively, since these concentrations exhibited the lowest Ct values compared with other concentrations (Tables 5 and 6).

**Table 4.** Ct values of rabies N gene positive control in varying primer concentrations.

| Primer concentration<br>(μM) | Ct value |  |  | %<br>Detection | Mean Cq | Cq SD |
| --- | --- | --- | --- | --- | --- | --- |
|  | R1 | R2 | R3 |  |  |  |
| 2.0 | 17.07 | 16.25 | 17.07 | 100 | 16.80 | 0.47 |
| 4.0 | 16.03 | 16.27 | 16.28 | 100 | 16.91 | 0.14 |
| 6.0 | 16.04 | 15.98 | 16.08 | 100 | 16.33 | 0.05 |
| 8.0 | 16.92 | 16.96 | 17.03 | 100 | 16.97 | 0.06 |
| 10.0 | 17.04 | 17.11 | 16.81 | 100 | 16.99 | 0.15 |

**Table 5.** Ct values of Rabies N gene positive control in varying probe concentrations.

| Probe concentration (in μM) | Ct value |  |
| --- | --- | --- |
|  | Positive control | No template control |
| 0.5 | 20.02 | NA |
| 1.0 | 18.98 | NA |
| 2.0 | 18.12 | NA |
| 3.0 | 17.63 | NA |

**Table 6.** Ct values of Rabies N gene positive control in varying annealing temperature.

| Annealing temperature (°C) | Ct value |
| --- | --- |
| 64.8 | 18.60 |
| 64.4 | 18.41 |
| 63.5 | 18.38 |
| 61.9 | 18.27 |
| 60.0 | 17.97 |
| 58.4 | 17.96 |
| 57.3 | 18.09 |
| 56.8 | 18.15 |
| NTC | NA |

To test for the stability of the primers and probe when prepared in larger volumes (larger sample number), a positive control was run against 20 no template controls. A single amplification (Ct= 17.63) was observed as exhibited by the positive control (Figure 1).

**Figure 1.**
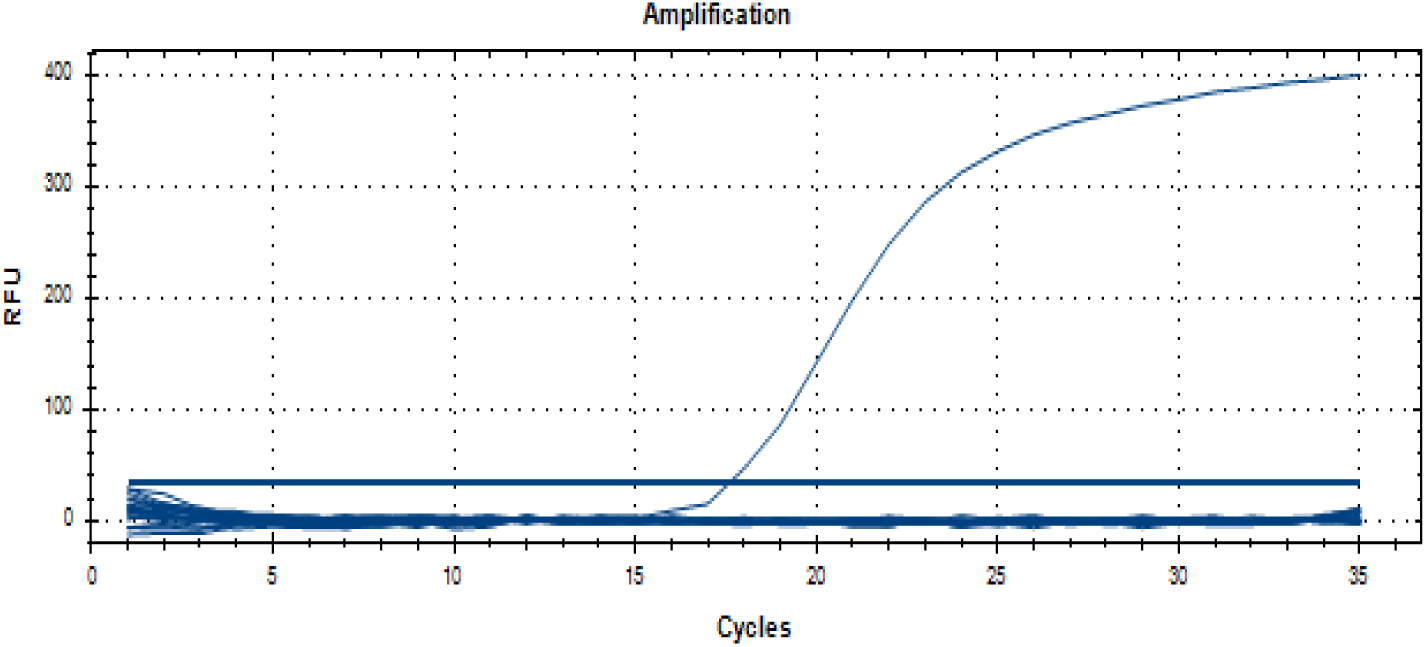
Amplification curve of rabies N gene positive control against 20 no template controls (NTC).

Next examined was the specificity of the assay to rule out non-specific amplification and detection. The cycle number for denaturation, annealing and extension was adjusted from 40 to 35 to remove possible non-specific amplification. The assay detected the rabies positive control samples while non-rabies samples (canine distemper virus and canine parvovirus) and no template controls were not amplified (Table 7).

**Table 7.** Clinical specificity of the RT-qPCR assay.

| Viral sample | Ct value |  |  |
| --- | --- | --- | --- |
|  | Replicate 1 | Replicate 2 | Replicate 3 |
| Rabies virus (PC) | 20.21 | 20.19 | 20.35 |
| Canine distemper virus | NA | NA | NA |
| Canine parvovirus | NA | NA | NA |
| No template control | NA | NA | NA |

Using 10-fold dilutions of the rabies viral RNA, a standard curve was established to determine the assay sensitivity. The established standard curve shows that the assay has a 95.1% efficiency (Figure 2). The limit of detection was determined by identifying the lowest standard dilution with at least 7/8 amplification (95% level of confidence). Results show that the assay can detect up to 1 RNA copy number/ul (Table 8).

**Figure 2.**
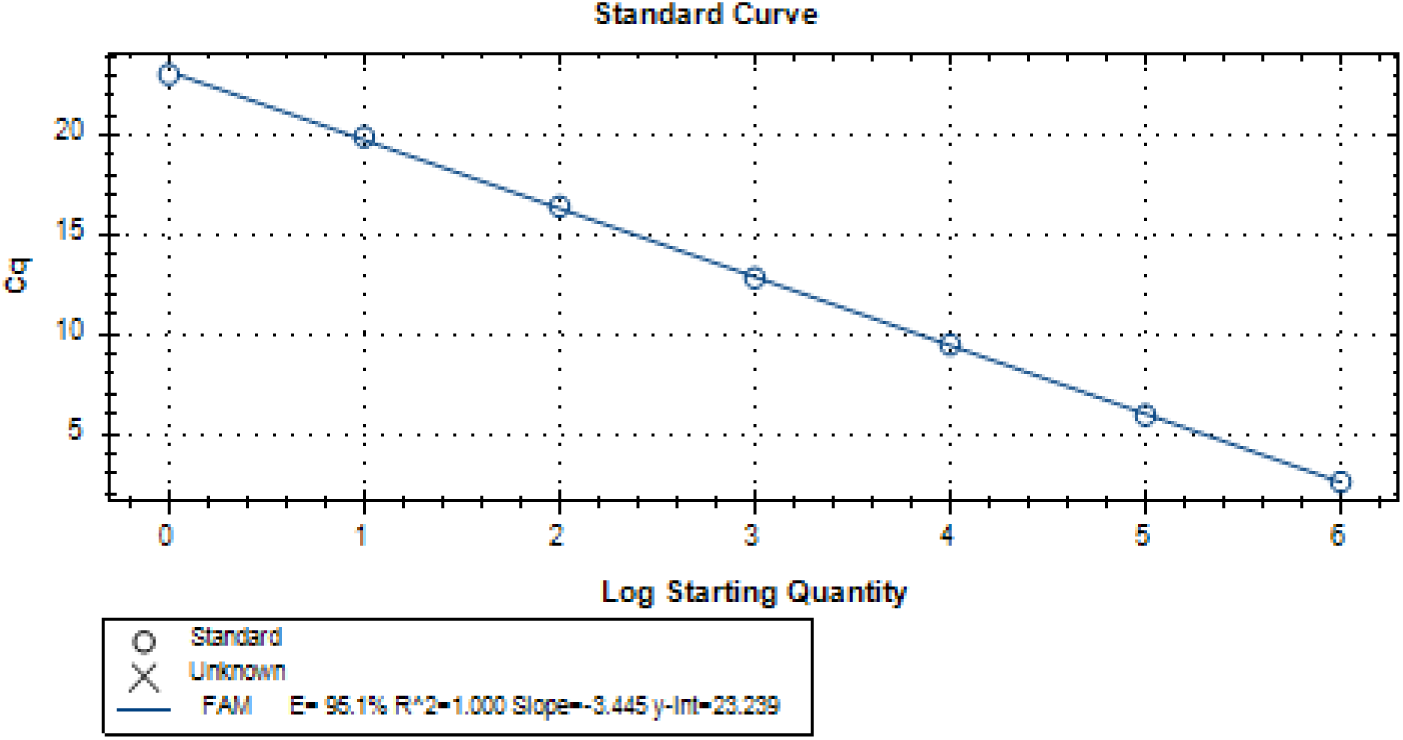
Standard curve of Rabies N gene.

**Table 8.** Determination of the assay limit of detection in different dilutions of rabies N gene positive control (8 replicates, R)

| RNA copy number /ul | Ct value |  |  |  |  |  |  |  |
| --- | --- | --- | --- | --- | --- | --- | --- | --- |
|  | R1 | R2 | R3 | R4 | R5 | R6 | R7 | R8 |
| 10<sup>6</sup> | 13.23 | 12.59 | 12.73 | 12.66 | 12.46 | 12.33 | 12.39 | 12.81 |
| 10<sup>4</sup> | 19.58 | 18.93 | 19.53 | 19.41 | 19.30 | 19.15 | 19.27 | 19.23 |
| 10<sup>3</sup> | 23.18 | 22.25 | 22.20 | 22.58 | 22.61 | 22.88 | 22.83 | 27.31 |
| 10<sup>2</sup> | 25.56 | 25.73 | 25.74 | 25.84 | 26.19 | 25.90 | 26.01 | 25.72 |
| 10<sup>1</sup> | 28.56 | 29.01 | 29.07 | 28.96 | 29.03 | 29.37 | 28.88 | 28.31 |
| 10<sup>0</sup> | 28.03 | 28.09 | 27.77 | 28.64 | 28.47 | 28.05 | 28.44 | 28.62 |
| 10<sup>-1</sup> | 28.93 | 29.18 | 29.98 | 29.39 | 30.62 | 28.85 | NA | NA |
| 10<sup>-2</sup> | 29.66 | 28.57 | 31.14 | 29.65 | 31.19 | 29.33 | NA | NA |
| 10<sup>-3</sup> | 30.81 | 31.28 | 31.62 | 30.46 | NA | NA | NA | NA |
| NTC | NA | NA | NA | NA | NA | NA | NA | NA |

To test reproducibility, a consistent result must be obtained when the assay is performed by different personnel using the same protocol and reagents within 1 week after the optimization. Results showed that the assay worked consistently even when performed by different personnel and different qPCR machines. No observable discrepancies were reported.

The optimized qPCR assay was validated with DFAT as the reference assay. Validation was done by extracting RNA from brain and non-brain samples (nasal planum and follicle sinus complexes) collected from DFAT-positive dogs and detecting the rabies viral RNA using the optimized assay. Table 9 shows that rabies N gene was detected from all brain and non-brain samples. Furthermore, only 27 of the 30 DFAT negative brain samples showed concordance to the DFAT results (Table 10).

**Table 9.** Detection of rabies N gene in DFAT positive brain and its corresponding nasal planum and follicle sinus complexes.

| Sample ID | Ct value |  |  |
| --- | --- | --- | --- |
|  | Brain | Follicle Sinus Complex | Nasal Planum |
| RADDL3-22-0219 | 27.04 | 29.14 | 26.80 |
| RADDL3-22-L097 | 12.70 | 28.68 | 27.22 |
| RADDL3-22-0112 | 20.40 | 26.23 | 26.11 |
| RADDL5-21-136 | 14.33 | 24.57 | 22.52 |
| RADDL5-21-140 | 16.70 | 26.31 | 27.22 |
| RADDL5-22-008 | 20.95 | 26.06 | 27.14 |
| RADDL6-21-4342 | 17.33 | 27.96 | 27.17 |
| RADDL6-21-4343 | 22.10 | 24.94 | 26.18 |
| RADDL6-21-4375 | 21.34 | 27.09 | 25.69 |
| RADDL7-21-149 | 15.48 | 24.69 | 26.55 |
| RADDL7-21-161 | 22.03 | 27.86 | 25.23 |
| RADDL2-22-9074 | 16.12 | 26.25 | 23.27 |
| RADDL2-22-5813 | 22.67 | 28.10 | 30.10 |
| RADDL2-22-9034 | 17.45 | 26.46 | 26.58 |
| RADDL11-22-001 | 18.82 | 25.91 | 18.20 |
| RADDL11-22-002 | 19.72 | 26.26 | 22.19 |
| RADDL4A-22-171 | 21.88 | 25.06 | 25.81 |
| RADDL4A-22-203 | 19.90 | 22.37 | 18.45 |
| CAR-22-020 | 18.83 | 24.88 | 24.08 |
| CAR-22-018 | 19.28 | 25.75 | 22.70 |

**Table 10.** Detection of rabies N gene in DFAT negative brain samples.

| Sample ID | Ct value | Sample ID | Ct value |
| --- | --- | --- | --- |
| Z-22-014 | NA | Z-22-093 | NA |
| Z-22-015 | NA | Z-22-095 | NA |
| Z-22-018 | NA | Z-22-097 | NA |
| Z-22-042 | NA | Z-22-098 | NA |
| Z-22-043 | NA | Z-22-099 | NA |
| Z-22-045 | NA | Z-22-106 | NA |
| Z-22-051 | NA | Z-22-109 | NA |
| Z-22-054 | NA | Z-22-114 | NA |
| Z-22-055 | NA | Z-22-117 | NA |
| Z-22-057 | NA | Z-22-022 | 30.46 |
| Z-22-059 | NA | Z-22-049 | 27.91 |
| Z-22-063 | NA | Z-22-081 | 32.95 |
| Z-22-064 | NA | Positive control 1 | 15.68 |
| Z-22-071 | NA | Positive control 2 | 15.67 |
| Z-22-076 | NA | Positive control 3 | 15.69 |
| Z-22-077 | NA | No template control 1 | NA |
| Z-22-079 | NA | No template control 2 | NA |
| Z-22-080 | NA | No template control 3 | NA |

## Discussion

The updated OIE Manual of Diagnostic Tests and Vaccines for terrestrial animals has recommended the use of RT-PCR assays for routine rabies diagnosis (WOAH, 2024). In this study we developed, optimized, and validated a real-time RT-PCR assay targeting the N gene for the detection of rabies virus strain in the Philippines. The nucleoprotein (N) gene is a highly conserved region critical for identifying genetic clusters related to rabies transmission in specific regions and species (Brunker et al., 2015; Troupin et al., 2016). Its sequences support large-scale epidemiological studies, pinpoint significant events, differentiate wildlife transmission cycles, and evaluate dog rabies elimination programs When monitoring is feasible, N-gene data enhances rabies surveillance and management, making it essential for understanding and improving rabies control efforts (Páez et al., 2007; Tao et al., 2009). Considering that brain samples can easily degrade when subjected to sub-optimal conditions (e.g. room temperature, prolonged transport, etc.), nasal planum and follicle sinus complex were used as alternate samples for assay validation.

During optimization, two temperatures 58.4°C and 60°C showed the two lowest Ct values (earlier amplifications), 17.96 and 17.97 respectively. However, it was observed that late non-specific amplification in non-rabies clinical samples is present when 58.4°C is used as the annealing temperature. (Hays et al., 2024; Rychlik et al., 1990). To prevent non-specific amplifications, the number of cycles for the amplification step was limited to 35 cycles only from an original 40 cycle setting. This change was critical for enhancing the accuracy of the assay. Increasing PCR cycle numbers can result in non-specific amplification, often caused by primer dimerization and excessive amplification of products. Primer dimers arise when primers bind to each other instead of the target DNA. Additionally, overamplification depletes reagents, leading to the formation of undesired products (Jansson & Hedman, 2019; Lorenz, 2012). The results confirmed that the assay detected rabies positive control samples only and none of the no template controls. Notably, non-rabies viral samples: canine distemper virus and canine parvovirus, and no-template controls, showed no amplification, demonstrating clinical specificity of the assay. These findings show that the assay can accurately distinguish rabies virus from other canine pathogens, indicating its potential as a valuable tool for clinical diagnostics and epidemiological monitoring.

A common method for validating qPCR assays involves establishing a standard curve, enabling the determination of the efficiency, linear dynamic range, and reproducibility of a qPCR assay. The efficiency of the assay should be 90–105%, and the R^2^ of the standard curve should be >0.980 (or r > –10.990) (Khairunisa et al., 2018). The assay developed has 95.2% efficiency and delivers comparable results regardless of the sample concentration or RNA present in the sample (Figure 2). The assay demonstrated an efficiency of 95.2% and an R² value exceeding 0.980, which indicates that it performs well within optimal ranges. PCR efficiency measures the amplification of the target sequence, with an ideal range being between 90% and 105%. The observed efficiency signifies minimal amplification bias, which is essential for accurately quantifying rabies viral RNA. The R² value indicates a strong linear relationship between template concentration and cycle threshold (Ct) values, allowing for dependable quantification across different RNA levels. This consistency makes the assay applicable in various clinical and research contexts, including situations where viral loads are low, such as in early infections or degraded samples. Additionally, the assay’s reliable performance across different sample qualities minimizes the risk of false results, enhancing its effectiveness for rabies diagnosis and monitoring, and ensuring accurate detection in a range of scenarios (Roe & Harnett, 2018; Khairunisa et al., 2018).

High assay sensitivity is crucial for the diagnosis of samples that have been transported or stored in less-than-ideal conditions and cannot be tested using other techniques. dFA testing requires cold-chain conditions that are not always feasible for sample transportation. Follicle sinus complexes and nasal planum, possible alternate specimens for rabies diagnosis (Shimatsu et al., 2016), were used for small-scale validation. All 60 (100%) samples showed concordance with the dFAT results, validating the assay’s capability in detecting rabies virus from non-brain samples, even at relatively low viral loads. Significant differences in the Ct value may be accounted for by the concentration of the rabies virus in each of the specimens extracted (Table 9). Muzzle skin or nasal planum and follicle sinus complexes typically have lower concentrations of the rabies virus compared to brain tissue because of how the virus behaves in the body.

The rabies virus primarily infects central nervous system (CNS), where it multiplies extensively in the brain and spinal cord. After a bite or other peripheral exposure, the virus travels through the nervous system to the CNS, where replication peaks, leading to higher viral loads in the brain (Jackson, 2013; Fooks et al., 2017). While muzzle skin and follicle sinus complexes are near the point of entry or peripheral nerves, they are not major sites for viral replication. The virus reaches these tissues later in the infection and in smaller amounts as it spreads outward from the CNS to peripheral areas like the salivary glands and skin. These tissues may contain the virus, but typically in much lower amounts compared to the brain, where replication is more intense (Wright et al., 2009; Zhang et al., 2014). Additionally, the skin and follicle sinus complexes have lower concentrations of nervous tissue and viral receptors than the brain, which reduces the amount of virus that accumulates in these areas (Shimatsu et al., 2016).

For the qPCR validation using dFAT-negative brain samples, there are three (3) samples which tested positive. The discrepancy in the results (3 of 30) may be accounted for by the low amounts of antigen present in the brain smear for dFAT. False-negative results in dFAT for rabies can arise when the concentration of viral antigens is too low. This is observed in the initial stages of infection, before the virus has fully reached the brain, or in late stages, when the viral load decreases as the infection spreads to other tissues. Additionally, testing areas of the brain with lower virus levels or using poorly preserved samples can lead to missed detection (Coetzer & Nel, 2019). However, these three samples may still have enough PCR-detectable viral RNA (Table 10).

In 2015, the WHO, OIE, FAO, and GARC have together set a goal of zero human dog-mediated deaths by 2030, globally. One of the objectives of this goal is to improve rabies surveillance in human and animals through improvement of diagnostics (WHO, OIE, FAO, and GARC, 2018). The developed qRT-PCR assay that has high throughput capability providing the potential to accelerate rabies testing in animals, and consequently enhance the rabies surveillance in the country. The assay can also be utilized in testing samples that are not suitable for dFA testing due to insufficient quantity or sample deterioration. Moreover, since the assay can be used for non-brain samples, it can alleviate the burden of sample collection, transport, and storage especially in resource-limited rural areas.

Although the assay shows promising results, several limitations should be acknowledged. It is currently limited to brain, nasal planum, and follicle sinus complexes, highlighting the need for further validation with additional non-brain samples to wider its scope. Furthermore, it requires verification with a larger sample size to confirm its robustness and applicability. The assay’s relatively high cost, attributed to its probe-based design, also limits its use to well-equipped laboratories; hence the recommendation to translate this to a cheaper qPCR detection like SYBR Green. Addressing these issues will be essential in increasing the assay’s utility in broader diagnostic and surveillance applications.

## Data Availability

Supplementary data that are not present in the manuscript are available and accessible upon request.

## Notes

### Competing Interest Statement

The authors have declared no competing interest.

### Funding Statement

The author(s) received no specific funding for this work.

### Author Declarations

Institutional Review Board (IRB) and Institutional Animal Care and Use Committee from Research Institute for Tropical Medicine

## REFEENCES

Centers for Disease Control and Prevention. How is rabies transmitted? [Internet]. 2019 [cited 2023 Feb 27]. Available from: https://www.cdc.gov/rabies/transmission/index.html

Centers for Disease Control and Prevention. Rabies [Internet]. 2021 [cited 2023 Feb 27]. Available from: https://www.cdc.gov/rabies/index.html

Department of Health. Rabies Prevention and Control Program [Internet]. 2020 [cited 2023 Feb 27]. Available from: https://doh.gov.ph/national-rabies-prevention-and-control-program

Domingo RD, Mananggit MR. Animal rabies patterns in Central Luzon, Philippines and implications for disease control. Philipp J Vet Med. 2014;51:117–24.

Duong V, Tarantola A, Ong S, Mey C, Choeung R, Ly S, et al. Laboratory diagnostics in dog-mediated rabies: an overview of performance and a proposed strategy for various settings. Int J Infect Dis. 2016;46:107–14. doi:10.1016/j.ijid.2016.03.016.

Mananggit MR, Kimitsuki K, Saito N, Garcia AMG, Lacanilao PMT, Ongtangco JT, et al. Background and descriptive features of rabies-suspected animals in Central Luzon, Philippines. Trop Med Health. 2021;49(1):59. doi:10.1186/s41182-021-00351.

Park CH, Kuboniwa S, Murakami R, Shiwa N, Inoue S, Kimitsuki K, et al. Immunohistochemical detection of virus antigen in the nasal planum of rabid dogs. J Vet Med Sci. 2021;83(10):1563–9. doi:10.1292/jvms.21-0438.

Ramírez GA, Rodríguez F, Herráez P, Suárez-Bonnet A, Andrada M, Espinosa-de-Los-Monteros A. Morphologic and immunohistochemical features of Merkel cells in the dog. Res Vet Sci. 2014;97(3):475–80. doi:10.1016/j.rvsc.2014.10.006.

Republic Act No. 9482. Anti-Rabies Act of 2007 (Philippines).

Rupprecht CE. Rhabdoviruses: Rabies Virus. In: Baron S, editor. Medical Microbiology. 4th ed. Galveston (TX): University of Texas Medical Branch at Galveston; 1996. Chapter 61.

Shimatsu T, Shinozaki H, Kimitsuki K, Shiwa N, Manalo DL, Perez RC, et al. Localization of the rabies virus antigen in Merkel cells in the follicle-sinus complexes of muzzle skins of rabid dogs. J Virol Methods. 2016;237:40–6. doi:10.1016/j.jviromet.2016.08.021.

Shiwa N, Nakajima C, Kimitsuki K, Manalo DL, Noguchi A, Inoue S, et al. Follicle sinus complexes (FSCs) in muzzle skin as postmortem diagnostic material of rabid dogs. J Vet Med Sci. 2018;80(12):1818–21. doi:10.1292/jvms.18-0519.

Shiwa N, Manalo DL, Boldbaatar B, Noguchi A, Inoue S, Park CH. Follicle-sinus complexes in muzzle skin of domestic and wild animals as diagnostic material for detection of rabies. J Vet Med Sci. 2020;82(8):1204–8. doi:10.1292/jvms.20-0252.

World Health Organization. Bulletin – Europe: Rabies [Internet]. [cited 2023 Feb 27]. Available from: https://www.who-rabies-bulletin.org/site-page/diagnosis-rabies

World Organization for Animal Health. RABIES. OIE Terrestrial Manual, Chapter 2.1.13 [Internet]. 2008 [cited 2023 Feb 27]. Available from: https://www.oie.int/fileadmin/Home/eng/Health_standards/tahm/pdf/2.01.13_RABIES.pdf

World Health Organization. Laboratory Techniques in Rabies. 5th ed., vol 1. Rupprecht CE, Fooks AR, Abela-Rider B, editors. 2018. ISBN: 978-92-4-151515-3.

World Health Organization. Rabies Fact Sheet [Internet]. 2021 [cited 2023 Feb 27]. Available from: https://www.who.int/en/news-room/fact-sheets/detail/rabies

Tobiume M, Sato Y, Katano H, Nakajima N, Tanaka K, Noguchi A, et al. Rabies virus dissemination in neural tissues of autopsy cases due to rabies imported into Japan from the Philippines: Immunohistochemistry. Pathol Int. 2009;59(8):555–66. doi:10.1111/j.1440-1827.2009.02406.x.

Mattner F, Henke-Gendo C, Martens A, Drosten C, Schulz TF, Heim A, et al. Risk of rabies infection and adverse effects of postexposure prophylaxis in healthcare workers and other patient contacts exposed to a rabies virus-infected lung transplant recipient. Infect Control Hosp Epidemiol. 2007;28(5):513–8. doi:10.1086/513614.

Maier T, Schwarting A, Mauer D, Ross RS, Martens A, Kliem V, et al. Management and outcomes after multiple corneal and solid organ transplantations from a donor infected with rabies virus. Clin Infect Dis. 2010;50(8):1112–9.

Picard-Meyer E, Bruyere V, Barrat J, Tissot E, Barrat MJ, Cliquet F. Development of a hemi-nested RT-PCR method for the specific determination of European Bat Lyssavirus 1. Comparison with other rabies diagnostic methods. Vaccine. 2004;22(15–16):1921–9. doi:10.1016/j.vaccine.2003.11.01.

Rychlik W, Spencer WJ, Rhoads RP. Optimization of the annealing temperature for DNA amplification in vitro. Nucleic Acids Res. 1990;18(21):6409–12. doi:10.1093/nar/18.21.6409.

Hampson K, Coudeville L, Lembo T, Sambo M, Kieffer A, Attlan M, et al. Estimating the Global Burden of Endemic Canine Rabies. PLoS Negl Trop Dis. 2015;9(4) doi:10.1371/journal.pntd.0003709.

Vos A, Freuling CM, Ortmann S, Kretzschmar A, Mayer D, Schliephake A, et al. An assessment of shedding with the oral rabies virus vaccine strain SPBN GASGAS in target and non-target species. Vaccine. 2018;36(6):811–7. doi:10.1016/j.vaccine.2017.12.076.

Perez AL. Antemortem diagnosis of rabies via Nuchal skin biopsy. Arch Dermatol. 2007;143(5):659. doi:10.1001/archderm.143.5.663-a.

Cullinane A, Garvey M. A review of diagnostic tests recommended by the World Organisation for Animal Health Manual of Diagnostic Tests and Vaccines for Terrestrial Animals. Rev Sci Tech. 2021;40(1). doi:10.20506/rst.40.1.3209.

Gigante CM, Dettinger L, Powell J, Seiders M, Condori RE, Griesser R, et al. Multi-site evaluation of the LN34 pan-lyssavirus real-time RT-PCR assay for post-mortem rabies diagnostics. PLoS One. 2018;13(5) doi:10.1371/journal.pone.0197074.

Wacharapluesadee S, Phumesin P, Supavonwong P, Khawplod P, Intarut N, Hemachudha T. Comparative detection of rabies RNA by NASBA, real-time PCR and conventional PCR. J Virol Methods. 2011;175(2):278–82. doi:10.1016/j.jviromet.2011.05.007.

McElhinney LM, Marston DA, Brookes SM, Fooks AR. Effects of carcass decomposition on rabies virus infectivity and detection. J Virol Methods. 2014;207:110–3. doi:10.1016/j.jviromet.2014.06.024.

David D, Yakobson B, Rotenberg D, Dveres N, Davidson I, Stram Y. Rabies virus detection by RT-PCR in decomposed naturally infected brains. Vet Microbiol. 2002;87(2):111–8. doi:10.1016/s0378-1135(02)00053-5.

Gilbert AT, Petersen BW, Recuenco S, Niezgoda M, Gómez JA, Laguna-Torres VA, et al. Evidence of rabies virus exposure among humans in the Peruvian Amazon. Am J Trop Med Hyg. 2012;87(2):206–15. doi:10.4269/ajtmh.2012.11-0689.

Dean DJ, Abelseth MK, Atanasiu P. The fluorescent antibody test. In: Meslin FX, Kaplan MM, Koprowski H, editors. Laboratory techniques in rabies. 4th ed. Geneva: WHO; 1996. p. 88–95.

